# Using Machine-Learning for Prediction of the Response to Cardiac Resynchronization Therapy: the SMART-AV Study

**DOI:** 10.1101/2020.07.16.20155424

**Authors:** Stacey J. Howell, Tim Stivland, Kenneth Stein, Kenneth A. Ellenbogen, Larisa G. Tereshchenko

**Author notes:** Correspondence: Larisa Tereshchenko, 3181 SW Sam Jackson Park Rd; UHN62; Portland, OR, 97239.

## Abstract

**Background:** There is a controversy whether the response of both sexes to cardiac resynchronization therapy (CRT) is similar. Optimal CRT delivery requires procedure planning.

**Objective:** To apply machine learning (ML) to develop a prediction model for CRT response.

**Methods:** Participants from the SmartDelay Determined AV Optimization (SMART-AV) trial (n=741; age, 66 ±11 yrs; 33% female; 100% NYHA III-IV; 100% EF≤35%) were randomly split into training & testing (80%; n=593), and validation (20%; n=148) samples. The entropy balancing procedure was used to match for the means of 30 covariates in male and female groups. Baseline clinical, ECG, echocardiographic and biomarker characteristics, and left ventricular (LV) lead position (43 variables) were included in 6 ML models (random forests, convolutional neural network, lasso, adaptive lasso, plugin lasso, elastic net, ridge, and logistic regression). A composite of freedom from death and heart failure hospitalization and a >15% reduction in LV end-systolic volume index at 6-months post-CRT was the endpoint.

**Results:** The primary endpoint was met by 337 patients (45.5%). Weighting resulted in a perfect balance of means of covariates in men and women. After reweighting, CRT response for women versus men was similar (OR 1.53; 95%CI 0.88-2.65; P=0.131). The adaptive lasso model was more accurate than class I ACC/AHA guidelines criteria (AUC 0.759; 95%CI 0.678-0.840 versus 0.639; 95%CI 0.554-0.722; *P*<0.0001), well-calibrated, and parsimonious (19 predictors; nearly half are potentially modifiable).

**Conclusions:** After balancing for covariates, both sexes similarly benefit from CRT. ML predicts short-term CRT response and thus may help with CRT procedure planning.

## Introduction

Cardiac resynchronization therapy (CRT) is an established treatment for patients with systolic heart failure (HF) and ventricular dyssynchrony.^1^ However, despite proven benefit, nearly a third of CRT recipients are considered to be “non-responders”.^2^ Furthermore, although many previous studies have suggested that female sex is associated with a higher responder rate, there is still controversy: some studies determined that the response of both sexes to CRT is similar.^3-5^

Guided left ventricular (LV) lead placement considering the timing of LV activation and electrical delay^6^, together with dynamic atrioventricular (AV) optimization^7^, can potentially reduce the CRT non-response rate. Previous analysis of the SMART-AV (SmartDelay Determined AV Optimization: A Comparison to Other AV Delay Methods Used in Cardiac Resynchronization Therapy) study showed an enhanced response to AV optimization in women as compared to men.^8^ Furthermore, the SMART-AV study suggested a strategy for using measures of LV electrical delay at implantation to guide LV lead placement.^9^ However, a complex interaction between cardiac veins anatomy and cardiomyopathy substrate can make guided LV lead placement procedure technically difficult. Prediction of the probability of a CRT response can possibly help with the allocation of resources and CRT procedure planning.

Machine learning (ML) has taken hold in a number of fields to improve risk prediction as compared to traditional methods.^10^ Several studies have applied ML to address the clinical challenge of CRT patient selection and showed that ML algorithms perform better than guidelines-recommended QRS duration and bundle branch block (BBB) morphology.^11-14^ However, all previous ML-prediction models targeted the long-term (≥ 1 year) CRT outcomes. At present, there is no short-term (6-month) CRT response prediction tool that can be used to plan CRT implantation and delivery.

We conducted the current study with two goals: (1) determine whether the response of both sexes to CRT is similar, and (2) use ML to predict short-term (6-month) response to CRT.

## Methods

The authors used the deidentified SMART-AV study dataset, provided by the executive study committee. The Oregon Health & Science University Institutional Review Board reviewed the current study and determined the deidentified nature of the dataset. We provided open-source code for statistical data analysis at https://github.com/Tereshchenkolab/statistics.

### Study population

The SMART-AV was a randomized, multicenter, single-blinded clinical trial^15, 16^ that sought to determine whether AV delay optimization would improve CRT response at six months post-implant. The trial enrolled New York Heart Association (NYHA) class III-IV HF patients with left ventricular ejection fraction (LVEF)≤35% despite optimal medical therapy, and QRS duration ≥120 ms, in sinus rhythm. HF patients who were in complete heart block, could not tolerate pacing at VVI-40-RV for up to two weeks, or previously received CRT were excluded. Enrollment was completed from May 2008 through December 2009. In the current study, we excluded participants with missing candidate predictor variables and lost to follow-up (Figure 1). Of the 980 randomized SMART-AV participants, 741 CRT recipients were included in this study.

**Figure 1:**
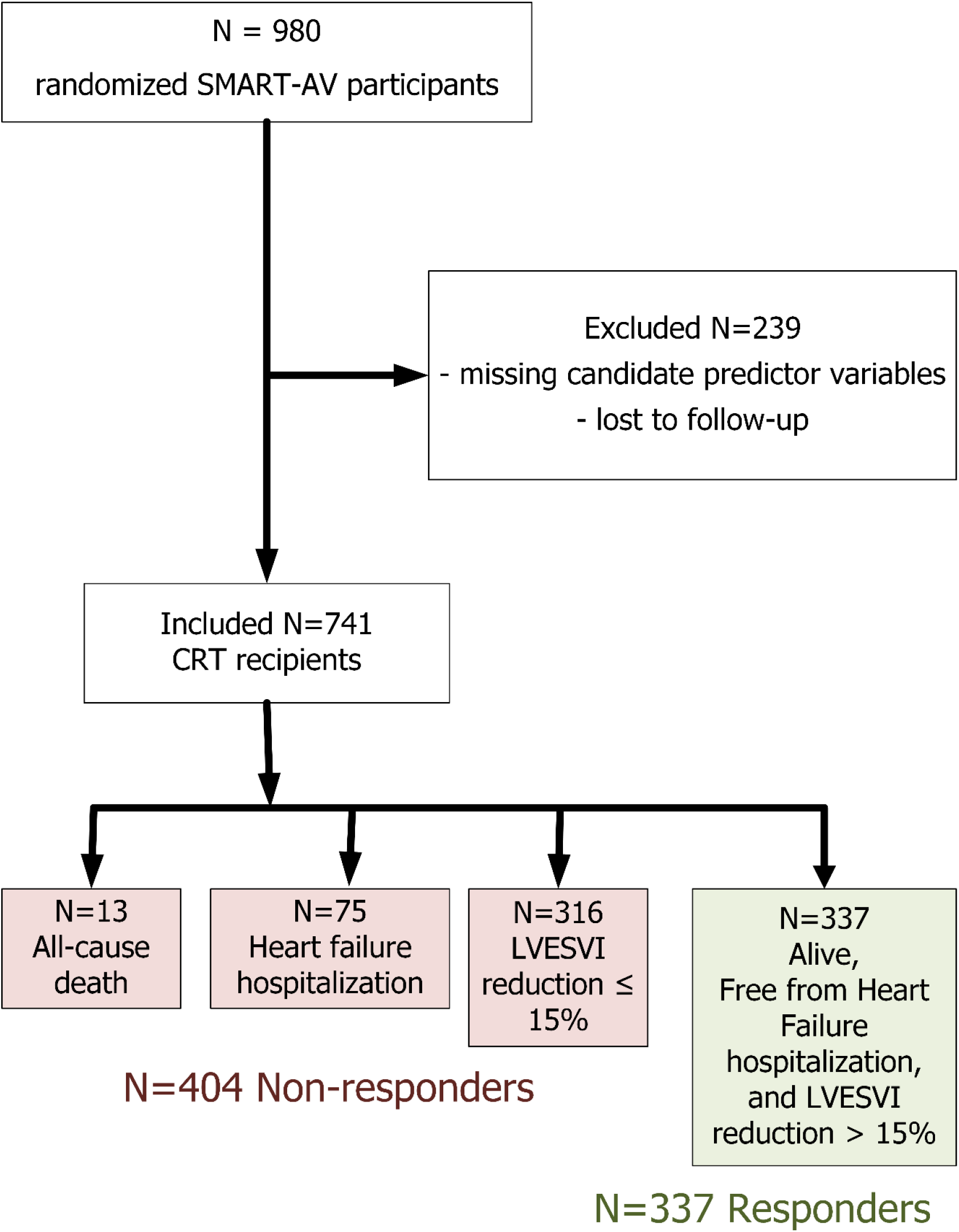
Flowchart of study cohort development.

### Candidate predictor variables

At the enrollment visit, baseline clinical characteristics data were collected, which included medical history, current cardiovascular evaluation (NYHA class) and medications list, the 6-minute walk test, quality of life (Minnesota Living with Heart Failure Questionnaire), and blood draw for biomarkers.^15, 16^ We calculated estimated glomerular filtration rate (eGFR) using the chronic kidney disease (CKD) Epidemiology Collaboration equation (CKD-EPI).^17^ LV lead location was selected at the discretion of the implanting physician. Baseline ECG and echocardiogram were recorded post-implant (no biventricular pacing).^15, 16^ We normalized LV volumes and dimensions by body surface area (BSA).

### The study endpoint

In the current study, we defined the primary endpoint as a composite of freedom from death and HF hospitalization and a >15% reduction^7-9, 18^ in LV end-systolic volume index (LVESVI) at six months of follow-up. LVESV was the primary endpoint in the SMART-AV trial.^15, 16^ A single core laboratory performed all echocardiographic measurements in a blinded fashion.

### Statistical analysis

#### Unadjusted comparisons

Normally distributed continuous variables were compared using the *t*-test and reported as mean ± standard deviation (SD). Variables with a skewed distribution were compared using the Wilcoxon rank-sum test and reported as the median and interquartile range (IQR). Categorical variables were compared using the χ2 test. Univariate logistic regression compared the odds of CRT response.

#### Adjusted analysis of sex differences in CRT response

Previous multivariate analysis of the SMART-AV data^8^ adjusted only for key sex differences in baseline characteristics. Previous regression analyses comparing CRT outcomes in men and women faced limitations due to the relatively small proportion of female participants and, therefore, were not able to conduct comprehensive adjustment for all known sex differences. To compensate for the noticeable difference in sex subgroup fractions and baseline characteristics of men and women in the study^8^, we used entropy balancing, a data matching procedure that allows reweighting a dataset such that the covariate distributions in the reweighted male group match the covariate moments in female group.^19^ We matched for the means of 30 covariates (Supplemental Table 1).

#### Prediction of the endpoint using machine learning

We randomly split the study population into two non-overlapping samples: training & testing (80%; n=593), and validation (20%; n=148). Considering future clinical implementation, we included routinely available predictor variables that describe baseline clinical, ECG, echocardiographic and biomarker characteristics, and LV lead position (43 variables, Table 1).

**Table 1.**
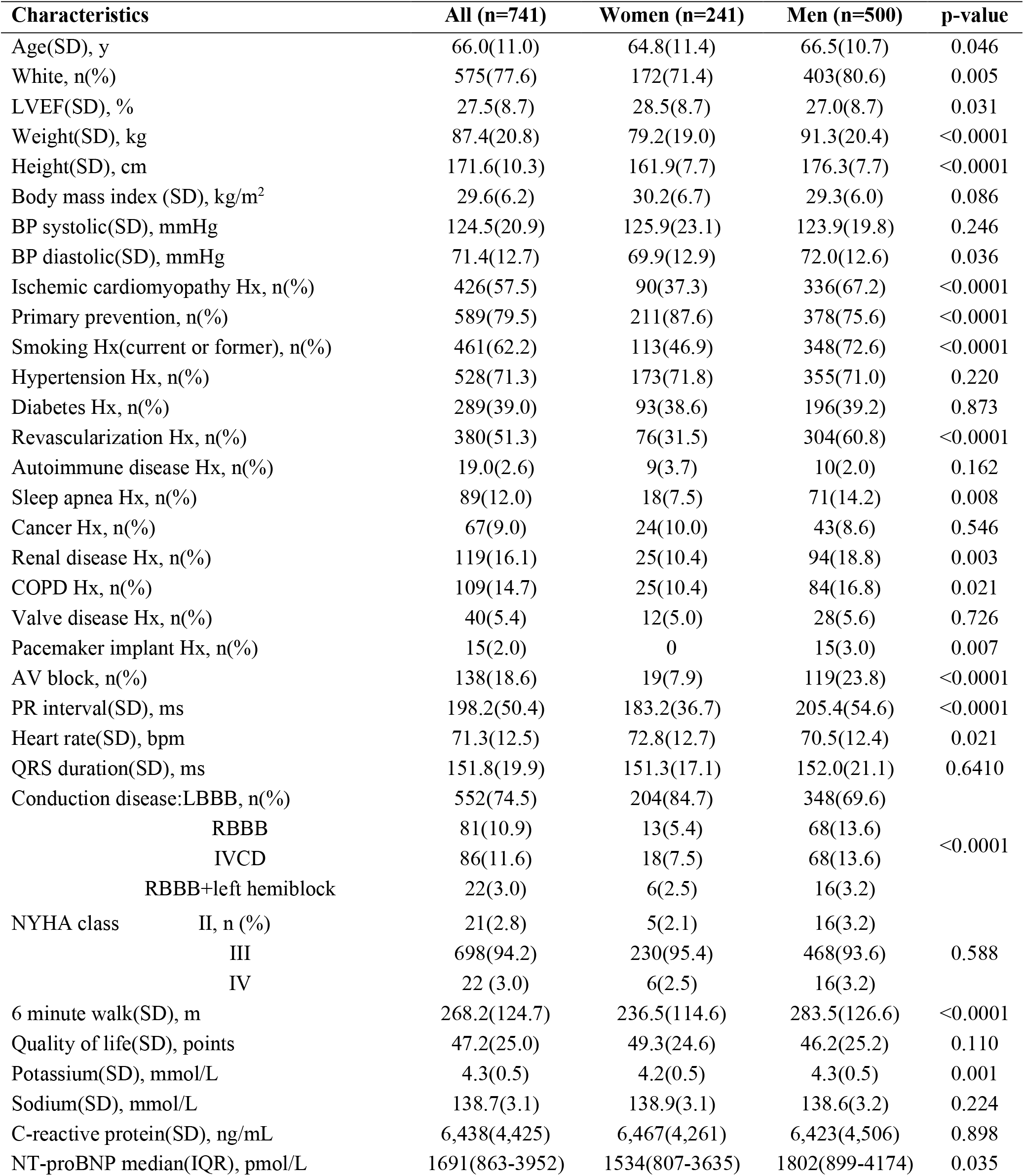
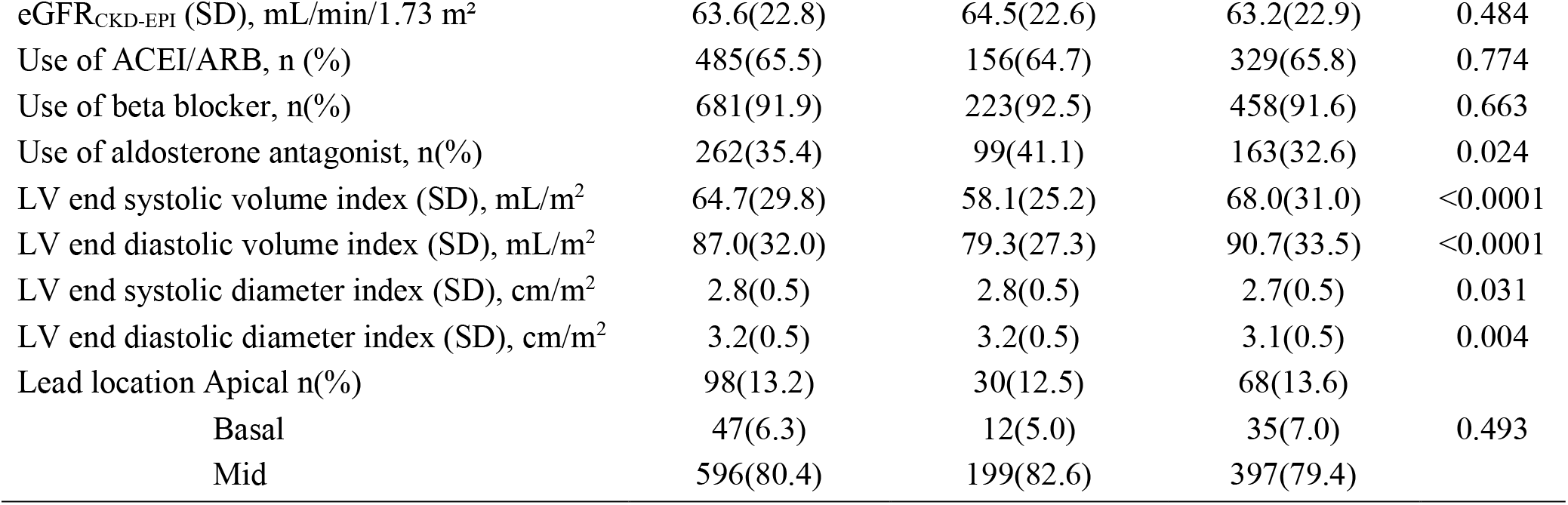
Baseline Clinical Characteristics in Men and Women.

We fitted eight different models (random forests^20^, convolutional neural network^21^, lasso, adaptive lasso, plugin lasso, elastic net, ridge, and logistic regression).

To train the random forests algorithm, we arranged the data in a randomly sorted order and tuned the number of subtrees and number of variables to randomly investigate at each split. We calculated both out-of-bag error (tested against training data subsets that are not included in subtree construction) and a validation error (tested against the validation data) to find the model with the highest testing accuracy.

We trained the convolutional neural network with 20 hidden layers, using 500 iterations with a training factor 2 and 4 normalization parameters. The network was comprised of 3 layers, 64 neurons per layer, and 901 synapse weights.

The family of lasso (least absolute shrinkage and selection operator) models employed ten-fold cross-validation in the training & testing sample. In lasso model, cross-validation selected the tuning parameter λ that minimized the out-of-sample deviance. The adaptive lasso performed multistep cross-validation, performing the second cross-validation step among the covariates selected in the first cross-validation step. The plugin lasso used partialing-out estimators to determine which covariates belong in the model, achieving an optimal bound on the number of covariates it included.^22^ The elastic net permitted retention of correlated covariates.^23^ In the ridge model, the penalty parameter used squared terms and kept all predictors in the model.

We validated the predictive accuracy of the models by comparing the area under the receiver operator curve (ROC AUC) in the validation sample. To assess calibration, we compared the observed and predicted proportions within the groups formed by the Hosmer-Lemeshow test^24^, and used the calibration belt^25^ to examine the relationship between out-of-sample estimated probabilities and observed CRT response rates. For the lasso family of models, we also calculated the out-of-sample deviance and deviance ratio.

We compared the performance of the selected model to the current 2013 American College of Cardiology Foundation/American Heart Association class I guideline criteria (QRS>150 ms and the presence of LBBB).^26^

Statistical analysis was performed using STATA MP 16.1 (StataCorp LP, College Station, TX). *P*-value < 0.05 was considered statistically significant.

## Results

### Study population

The SMART-AV study population characteristics in men and women are vastly different, as shown in Table 1 and have been previously reported elsewhere.^8^ At baseline, men were more likely white, with ischemic cardiomyopathy, AV block, a lower LVEF, and higher LVESVI, LVEDVI, and NT-proBNP. Women more likely had LBBB and used aldosterone antagonists. There was no difference in LV lead location between men and women.

### Comparison of the response of men and women to CRT

The primary endpoint was met by 337 patients (45.5%), 134 were women (55.6% response), and 203 were men (40.6% response); *P*<0.0001. Out of 404 participants who failed to respond, 13 died (10 men and 3 women), 75 participants (53 men and 22 women) were hospitalized because of HF, and 316 (334 men and 82 women) participants failed to achieve a volumetric response.

Univariate logistic regression analysis showed that women had an 80% higher probability for composite CRT response (Odds ratio (OR) 1.83; 95%CI 1.34-2.50); *P*<0.0001). Weighting resulted in a perfect balance of means of covariates in men and women (Supplemental Table 1). After reweighting, there was no significant difference in CRT response for women as compared to men (OR 1.53; 95%CI 0.88-2.65; *P*=0.131).

### Prediction of CRT response

In tuning the random forests algorithm, we observed that both out-of-bag error and validation error stabilized after 300 iterations at 30-35% (Supplemental Figure 1), and we conservatively chose 500 subtrees. The minimum validation error was observed for 7 variables, and we chose 7 variables to investigate at each split randomly. The final random forests model reported 26% error in validation sample; it accurately predicted freedom from composite CRT response endpoint in 71 out of 83 participants (specificity 85.5%), and correctly predicted CRT response in 38 out of 65 individuals (sensitivity 58.5%), having a positive predictive value of 76% and negative predictive value of 72.4%. The single most important predictor was diabetes (Figure 2), which, together with demographic characteristics (age, sex, race) and other comorbidities (hypertension, smoking) comprised six the most important predictors.

**Figure 2.**
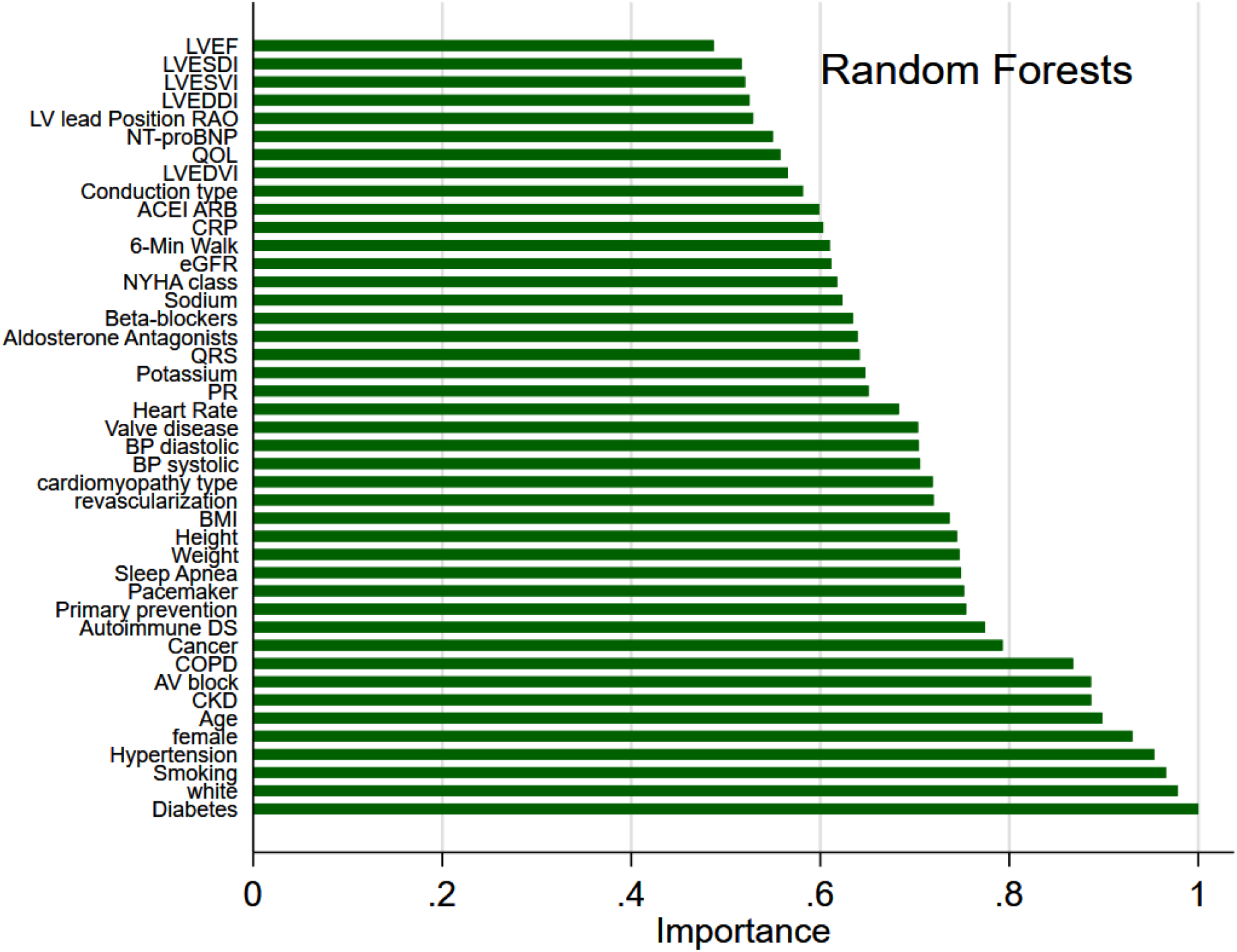
Importance scores of predictor variables in the random forests model.

A comparison of the prediction models’ performance is shown in Table 2. The convolutional neural network demonstrated the highest predictive accuracy in the training & testing sample, with a final error of only 6%. However, the calibration of the convolutional neural network model was unsatisfactory (Hosmer-Lemeshow test P<0.0001; Supplemental Figure 2), and predictive accuracy in the validation sample did not differ from the lasso family of models.

**Table 2.** Development and validation of composite CRT response prediction tool.

| Group | Model | Training & testing sample (N=593) |  |  |  |  | Validation sample (N=148) |  |  |  |  |
| --- | --- | --- | --- | --- | --- | --- | --- | --- | --- | --- | --- |
|  |  | Deviance | Deviance ratio | Number of predictors | ROC AUC (95%CI) | P-value | Deviance | Deviance ratio | N predictors | ROC AUC (95%CI) | P-value |
| All participants | Ridge | 1.201 | 0.129 | 43 | 0.753(0.714-0.792) |  | 1.164 | 0.151 | 43 | 0.778(0.699-0.857) |  |
|  | Elastic net | 1.196 | 0.133 | 30 | 0.751(0.711-0.790) |  | 1.163 | 0.152 | 30 | 0.769(0.688-0.849) |  |
|  | Lasso | 1.187 | 0.140 | 29 | 0.752(0.713-0.792) | 0.277 | 1.155 | 0.158 | 29 | 0.770(0.690-0.850) |  |
|  | Adaptive lasso | 1.184 | 0.142 | 19 | 0.751(0.712-0.790) |  | 1.169 | 0.148 | 19 | 0.759(0.678-0.840) | 0.368 |
|  | Logistic regress | 1.147 | 0.168 | 43 | 0.768(0.730-0.805) |  | 1.135 | 0.172 | 43 | 0.774(0.697-0.851) |  |
|  | CNN | - | - | 43 | 0.979(0.966-0.993) | <0.0001 | - | - | 43 | 0.759(0.682-0.837) |  |
|  | Random forest | - | - | 43 | 0.642(0.600-0.683) | <0.0001 | - | - | 43 | 0.720(0.649-0.791) |  |
|  | Plugin lasso | 1.295 | 0.061 | 2 | 0.655(0.613-0.696) | <0.0001 | 1.296 | 0.055 | 2 | 0.667(0.582-0.751) | 0.028 |
| All coefficients are penalized except plugin lasso (postselection) |  |  |  |  |  |  |  |  |  |  |  |

Several models (lasso, adaptive lasso, elastic net, ridge, and logistic regression) demonstrated similar fit and predictive accuracy both in training & testing, and validation samples (Table 2), which was significantly higher than current class I clinical guidelines (AUC 0.639; 95%CI 0.554-0.722), *P*<0.0001. Figure 3 shows the cross-validation function and selected λ for each model. Only a few models (logistic regression, adaptive lasso, and plugin lasso) showed satisfactory out-of-sample calibration (Figure 4). Ultimately, we selected the adaptive lasso model as the most accurate, well-calibrated, and parsimonious (19 predictors listed in Supplemental Table 2).

**Figure 3.**
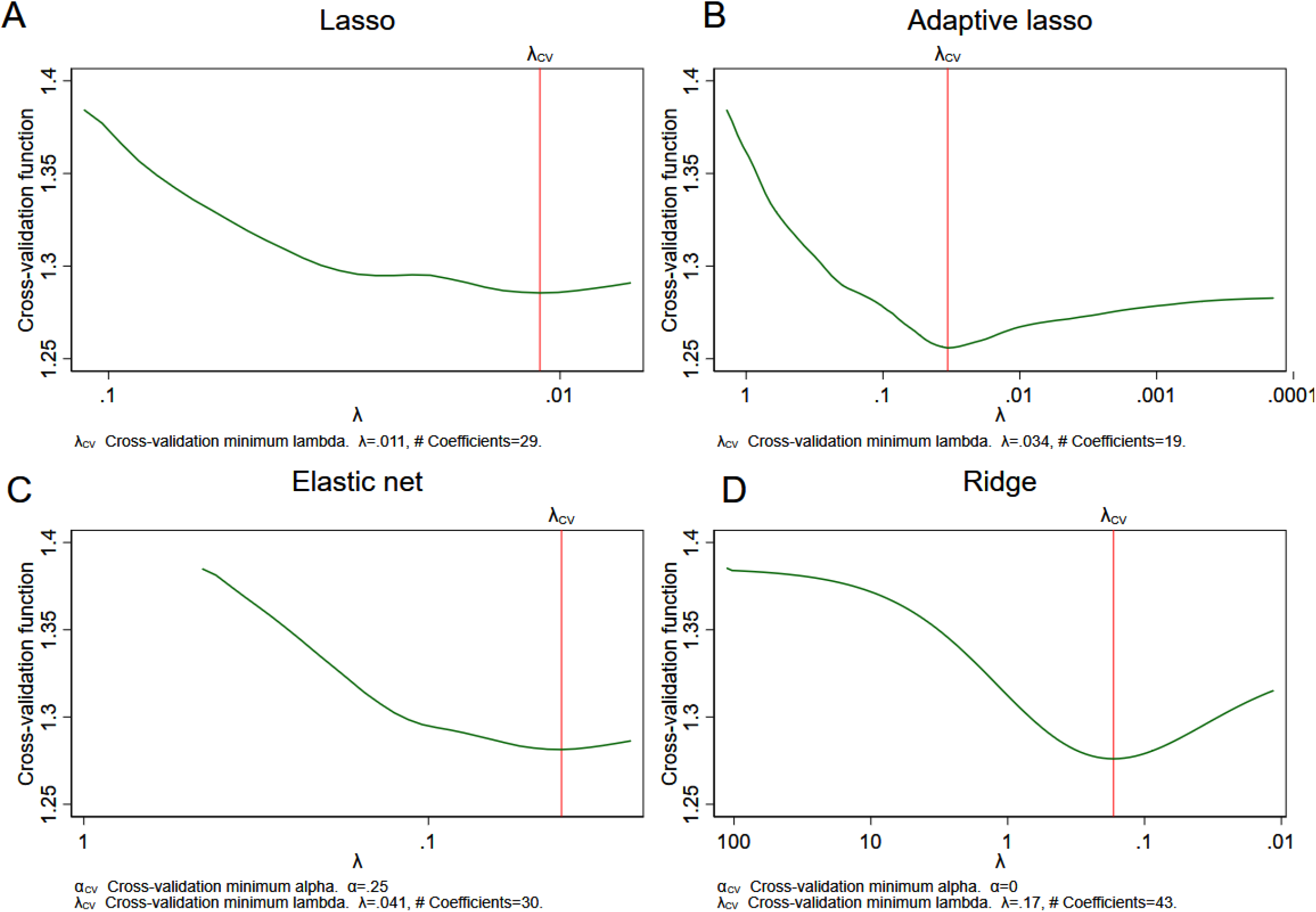
Cross-validation (CV) function (the mean deviance in the CV samples) is plotted over the search grid for the lasso penalty parameter λ on a reverse logarithmic scale for (**A**) lasso, (**B**) adaptive lasso, (**C**) elastic net, (**D**) ridge models. The first λ tried is on the left, and the last λ tried is on the right.

**Figure 4.**
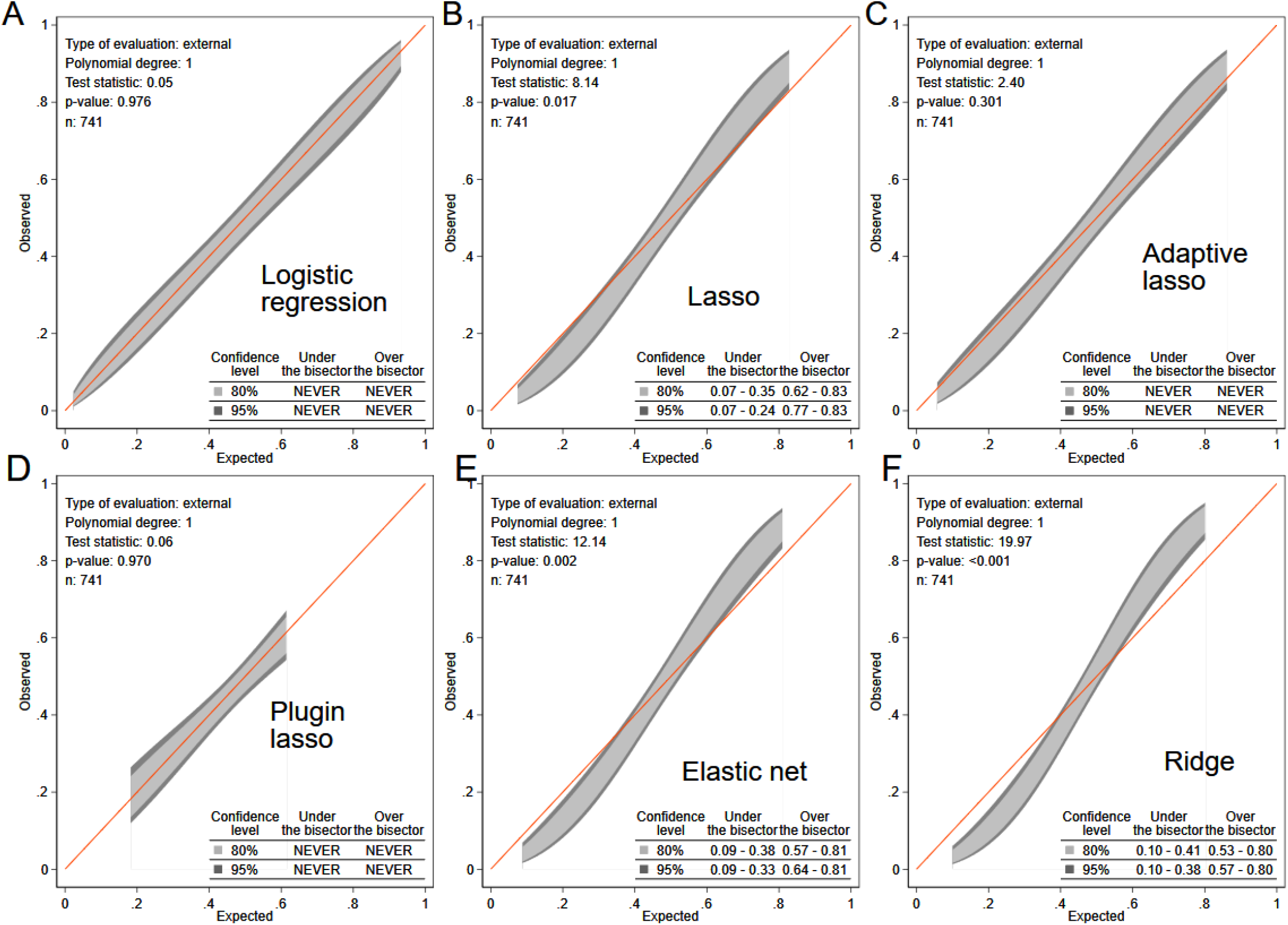
The calibration belt with 80% and 95% confidence intervals on the external sample shows the observed and predicted CRT response proportions in (**A**) logistic regression, (**B**) lasso, (**C**) adaptive lasso, (**D**) plugin lasso, (**E**) elastic net, and (**F**) ridge models for all participants.

In the adaptive lasso model, the most important predictors (Figure 5) characterized dyssynchrony (ventricular conduction type, QRS duration), underlying disease substrate (cardiomyopathy type, primary prevention indication), and modifiable characteristics (NT-proBNP, systolic blood pressure), including PR interval. Nonischemic cardiomyopathy, female sex, primary prevention indication, history of valvular heart disease and cancer, and higher QRS duration, systolic blood pressure, LVEDVI, 6-min walk distance, eGFRCKD-EPI, and age were associated with CRT response. Non-LBBB, AV block, and higher NT-proBNP, CRP, PR interval, LVEF, LVESDI, and weight were associated with non-response. Participants in the 5^th^ quantile as compared to those in the 1^st^ quantile had 14-fold higher odds of composite CRT response (Figure 6). The online calculator is freely available at www.ecgpredictscd.org

**Figure 5.**
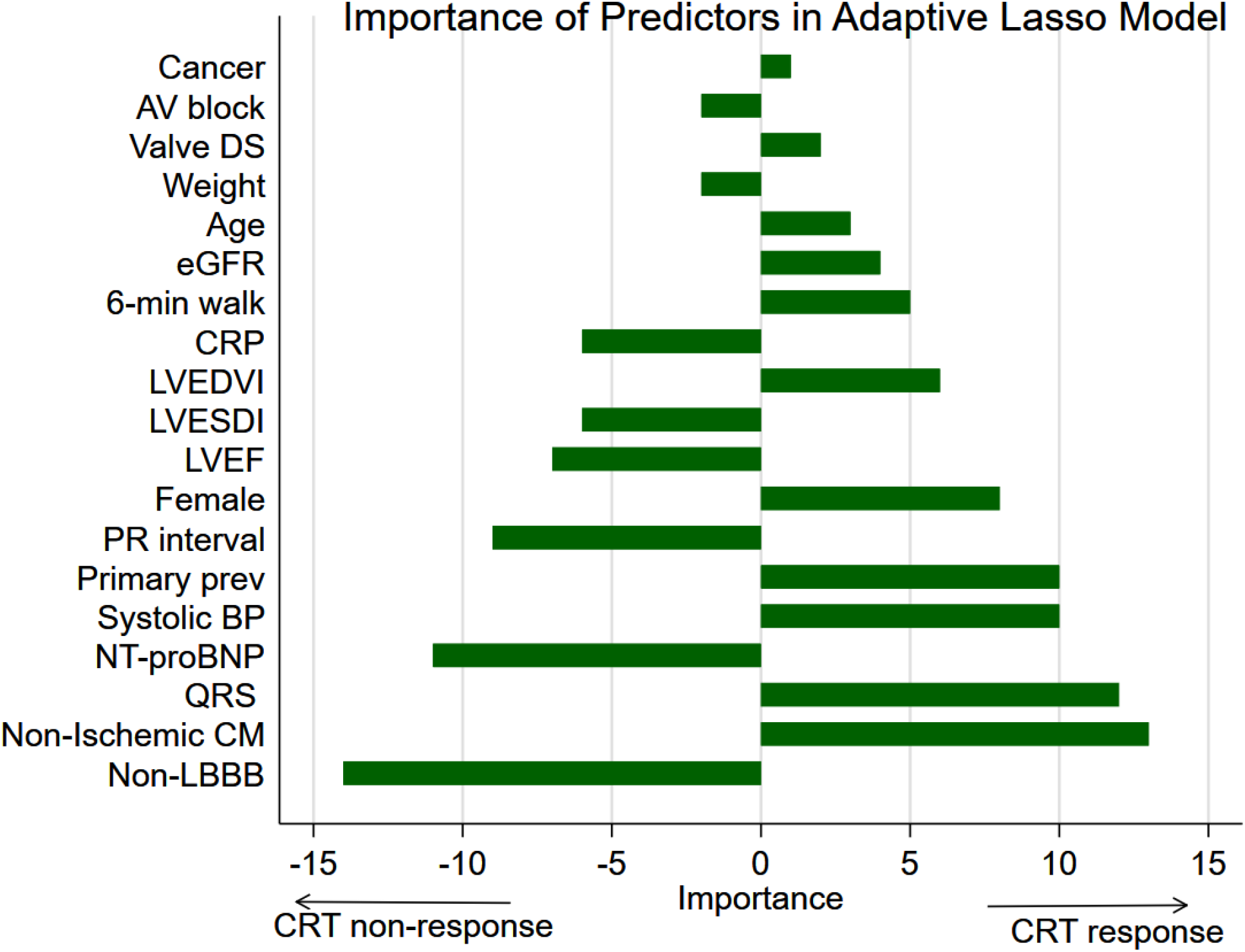
Importance of the selected predictors in the adaptive lasso model. The most important predictors were added to the model early.

**Figure 6.**
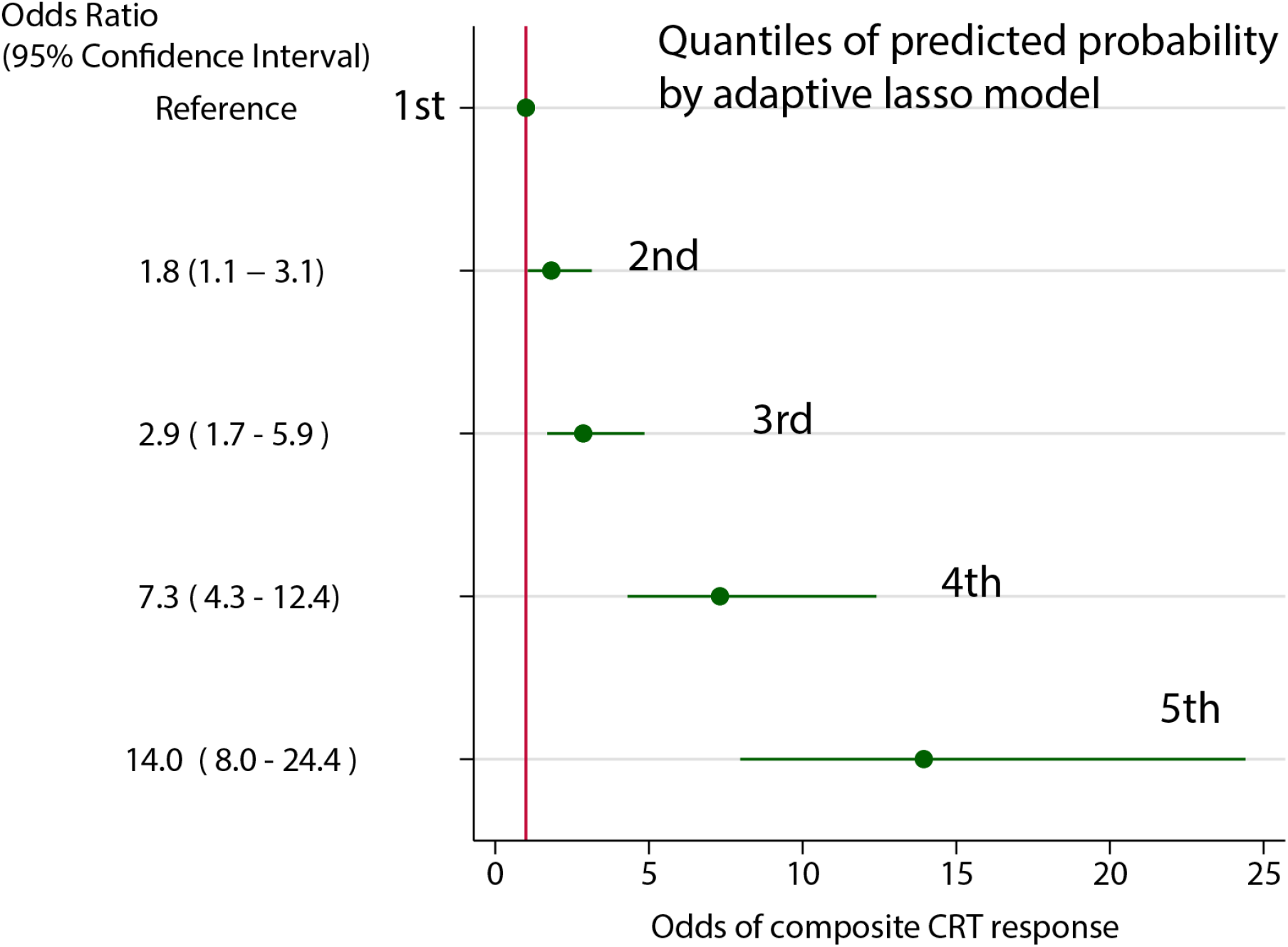
Probabilities of composite CRT response by quantiles of the adaptive lasso model.

## Discussion

There are two main findings in our study. First, we showed that both sexes equally benefit from CRT. Frequently observed better CRT outcomes in women than men are explained by the sex differences in baseline characteristics, including the disease substrate, dyssynchrony, comorbidities, and HF treatment. Second, using the ML approach, we developed a parsimonious model for the prediction of CRT response that is comprised of routinely available baseline clinical, ECG, and echocardiographic characteristics - measures of the disease substrate, dyssynchrony, and comorbidities. Importantly, many included predictors could be potentially modifiable. The model included both the PR interval and AV block, suggesting the importance of the dynamic AV optimization. Developed in this study, the short-term CRT response prediction model opens an avenue for a future randomized controlled trial, testing CRT implantation planning strategy, incorporating targeted lead placement and dynamic AV optimization programming.^7, 9^

### The response of both sexes to CRT is similar

Our study adds to the significant body of evidence indicating that fundamentally, men and women respond to CRT similarly.^3-5^ Men and women have substantial differences in underlying disease substrate, degree, and characteristics of dyssynchrony^3^, a spectrum of comorbidities, and HF treatment. Therefore, many previous studies concluded that CRT is more beneficial for women than men.^8^ Similar trends in the most predictive baseline comorbidities are seen in the landmark MADIT-CRT study, supporting the significance of our observation and the modeling done in the current study.^27, 28^ Importantly, our study utilized an appropriate analytical approach and demonstrated that both sexes benefit from CRT similarly. We used entropy balancing – the advanced matching analytical approach^19^ that efficiently balanced sex differences. Conclusion about the similar benefit for both sexes is important; it demystifies sex-specific CRT response and removes ground for sex inequality. However, our finding of similar benefit from CRT for both sexes does not negate sex differences in dyssynchrony, HF substrate, and response to pacing therapy^8, 29^, which have to be studied further.

### Prediction of composite CRT Response in a Short-Term – perspective for planning LV lead placement and CRT delivery

It has been previously shown that increasing degrees of interventricular (rather than intraventricular) dyssynchrony is expected to result in improved rates of clinical CRT response.^30^ Previous analysis of the SMART-AV study showed that optimally timed AV delay provides an incremental benefit to the substantial interventricular conduction delay^7, 9^, suggesting that both LV lead and right ventricular (RV) lead placement should target maximizing RV-LV delay. Pre-procedural planning may involve expensive and time-consuming cardiac imaging. Our risk score can predict the probability of the short-term composite CRT response and, therefore, can help to preserve resources while improving clinical outcomes. Careful pre-procedural planning would be particularly critical for CRT candidates with a moderate or low probability of CRT response, especially if they have modifiable factors. Notably, both the baseline PR interval and the presence of AV block were selected by the adaptive lasso model as essential predictors in the model, indicating the likely benefit of dynamic AV optimization.

Consistently with prior studies^9, 11-14^, we confirmed that ML could improve patient selection for CRT therapy beyond current guidelines. The strength of ML algorithms is the ability to capture complex interactions.^31^ Several prior studies have used ML to predict CRT response. Kalscheur et al analyzed 595 COMPANION NYHA III/IV patients,^11^ Cikes et al studied 1106 MADIT-CRT NYHA class ≤ II patients,^14^ Feeny et al evaluated 470 NYHA I-IV patients from an observational cohort, and Hu et al retrospectively analyzed 990 predominately NYHA II-III patients from a single-center cohort.^32^ Of note, all previous studies considered long-term CRT benefits, answering a question of CRT candidate selection. In contrast, our prediction model is focusing on a short-term CRT response and can help planning the CRT delivery strategy, in addition to selecting the most appropriate CRT candidate. Distinguishing those at high risk of non-response could alert cardiologists to a specific group that requires special attention within the first six months after CRT implantation.

Presently response and outcomes following CRT implantation vary significantly^2^, making it crucial to improve patient selection for CRT. Improved identification of CRT responders could help to avoid CRT implantation in patients unlikely to benefit and to disproportionately incur undue harm and risk. Better prediction of CRT non-responders could be used to identify patients that may be better served with earlier consideration of advanced HF therapies, including mechanical circulatory support and transplantation rather than CRT, which would carry a lower yield of clinical improvement.

In this study, an absence of sustained ventricular tachyarrhythmia (primary prevention indication) was an important predictor of CRT response. This finding is consistent with previous studies that showed the antiarrhythmic effect of CRT and reversed electrical remodeling^33^, which can be facilitated by the autonomic nervous system response.^34^

A comparison of ML models and selection of the “best” model also deserves discussion. We observed similar accuracy in all but one (plugin lasso) models, leaving seven models for consideration. However, only two of them (logistic regression and adaptive lasso) demonstrated satisfactory calibration. The parsimonious model (adaptive lasso) won because of (1) convenience (19 versus 43 predictors), and (2) approach to feature importance ranking. The most important predictors in the random forests model describe comorbidities and demographic characteristics, which unlikely to be modified (age, sex, race, diabetes, hypertension, smoking). In contrast, the most important predictors in the adaptive lasso model provide a meaningful characterization of the disease substrate and its electrophysiology (a type of cardiomyopathy and conduction abnormality, QRS duration, history of sustained ventricular tachyarrhythmia or cardiac arrest, NT-proBNP and systolic blood pressure), which can guide CRT delivery.

### Strengths and Limitations

SMART-AV is a large multicenter randomized control trial with careful phenotyping that included blinded analysis of echocardiograms and biomarkers in core laboratories, and appropriate follow-up, providing an opportunity to study composite CRT response. A strength of the present study was the use of a composite endpoint of clinical outcomes (death, HF hospitalization) and volumetric remodeling. However, limitations of the study have to be taken into account. The study population was predominantly men, although this is characteristic and similar to other CRT trials. We limited candidate predictor variables by currently widely available and did not include novel ECG measures of dyssynchrony that can potentially further improve prediction.^18, 35^

## Conclusion

The response of both sexes to CRT is similar, as outcome disparities between sex subgroups are substantially explained by differences in disease substrate, characteristics of dyssynchrony, comorbidities, and HF treatment. ML predicts short-term CRT response and thus may help with CRT procedure planning.

## Data Availability

We provided open-source code for statistical data analysis at https://github.com/Tereshchenkolab/statistics.

## Abbreviations

AV: atrioventricular
ACEI/ARB: angiotensin converting enzyme inhibitor or angiotensin II receptor blocker
CM: cardiomyopathy
CRT: cardiac resynchronization therapy
EF: ejection fraction
HF: heart failure
LBBB: left bundle branch block
LV: left ventricular
ML: machine learning
NYHA: New York Heart Association
SMART-AV: SmartDelay Determined Atrioventricular Optimization
RCT: randomized controlled trial
BSA: body surface area
OR: odds ratio

---

**Supplemental Table 1.** The balance table of means of covariates before and after weighting.

| Characteristics means | Before balancing |  | After balancing |  |
| --- | --- | --- | --- | --- |
|  | Women (n=241) | Men (n=500) | Women (n=241) | Men (n=500) |
| Age, y | 64.78 | 66.53 | 64.78 | 64.78 |
| White | 0.71 | 0.81 | 0.71 | 0.71 |
| LVEF | 0.29 | 0.27 | 0.29 | 0.29 |
| Body mass index, kg/m<sup>2</sup> | 30.18 | 29.30 | 30.18 | 30.18 |
| Nonischemic cardiomyopathy | 0.63 | 0.33 | 0.63 | 0.63 |
| Primary prevention | 0.88 | 0.76 | 0.88 | 0.88 |
| Never smoking | 0.53 | 0.30 | 0.53 | 0.53 |
| Hypertension | 0.72 | 0.71 | 0.72 | 0.72 |
| Diabetes | 0.39 | 0.39 | 0.39 | 0.39 |
| Revascularization | 0.32 | 0.61 | 0.32 | 0.32 |
| Sleep apnea | 0.07 | 0.14 | 0.07 | 0.07 |
| Cancer | 0.10 | 0.09 | 0.10 | 0.10 |
| Renal disease | 0.10 | 0.19 | 0.10 | 0.10 |
| COPD | 0.10 | 0.17 | 0.10 | 0.10 |
| Valve disease | 0.05 | 0.06 | 0.05 | 0.05 |
| AV block | 0.08 | 0.24 | 0.08 | 0.08 |
| PR interval, ms | 183.2 | 205.4 | 183.20 | 183.20 |
| QRS duration, ms | 151.3 | 152.0 | 151.3 | 151.3 |
| Conduction disease: IVCD | 0.075 | 0.136 | 0.075 | 0.075 |
| RBBB | 0.054 | 0.136 | 0.054 | 0.054 |
| NYHA class III | 0.95 | 0.94 | 0.95 | 0.95 |
| IV | 0.02 | 0.03 | 0.02 | 0.02 |
| 6 minute walk, m | 236.5 | 283.5 | 236.5 | 236.5 |
| Quality of life, points | 49.27 | 46.15 | 49.27 | 49.27 |
| eGFR<sub>CKD-EPI</sub>, mL/min/1.73 m<sup>2</sup> | 64.48 | 63.23 | 64.48 | 64.48 |
| Use of ACEI/ARB | 0.85 | 0.84 | 0.85 | 0.85 |
| Use of beta blocker | 0.93 | 0.92 | 0.93 | 0.93 |
| Use of aldosterone antagonist | 0.41 | 0.33 | 0.41 | 0.41 |
| LVESVI, mL/m<sup>2</sup> | 58.05 | 67.97 | 58.05 | 58.05 |
| LVEDVI, mL/m<sup>2</sup> | 79.35 | 90.70 | 79.35 | 79.35 |
| LVESDI, cm/m<sup>2</sup> | 2.81 | 2.73 | 2.81 | 2.81 |
| LVEDDI, cm/m<sup>2</sup> | 3.24 | 3.13 | 3.24 | 3.24 |

**Supplemental Table 2.**
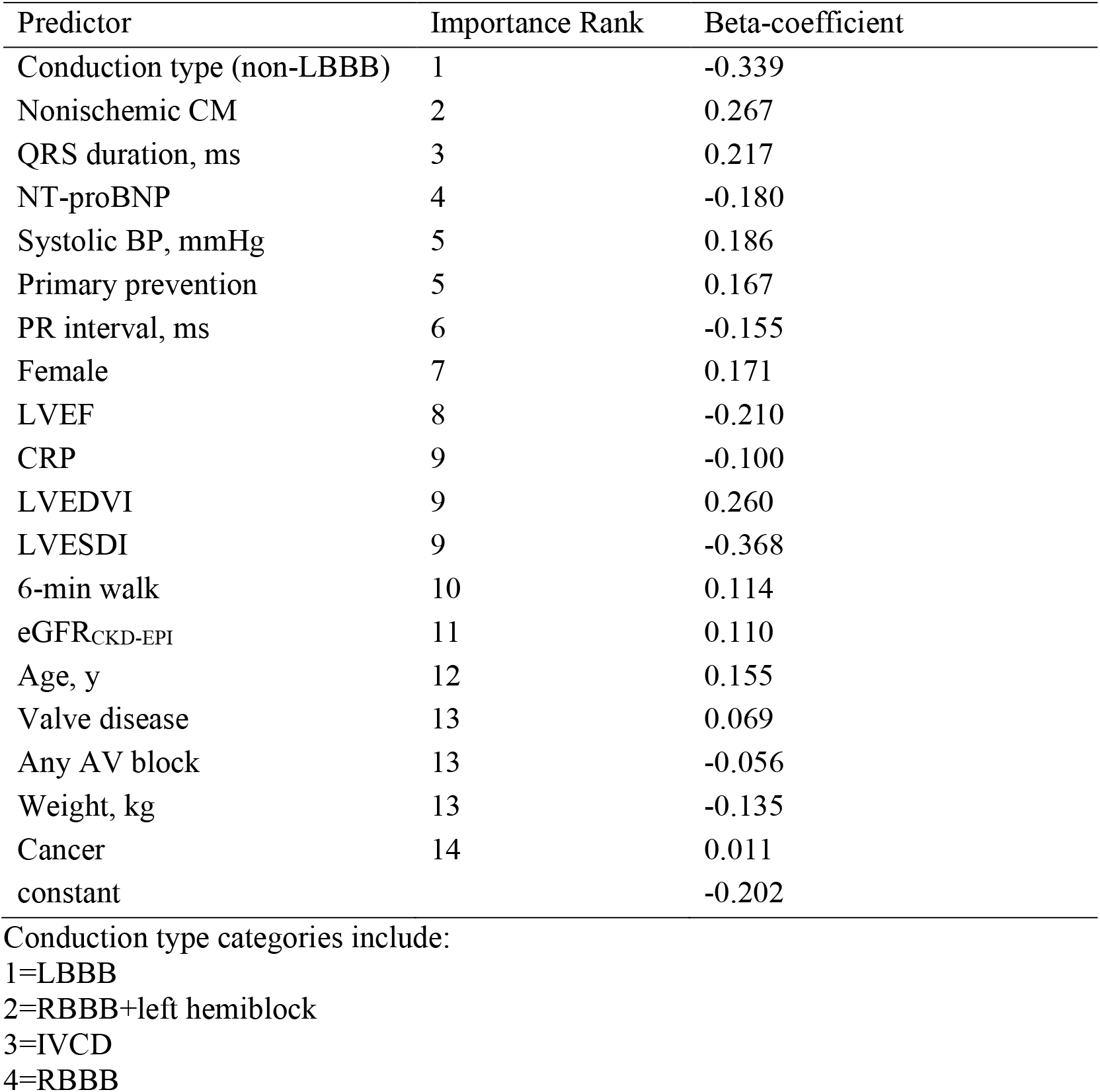
Lassoknots, or predictors in the adaptive lasso model listed in the order of their importance.

**Supplemental Figure 1.**
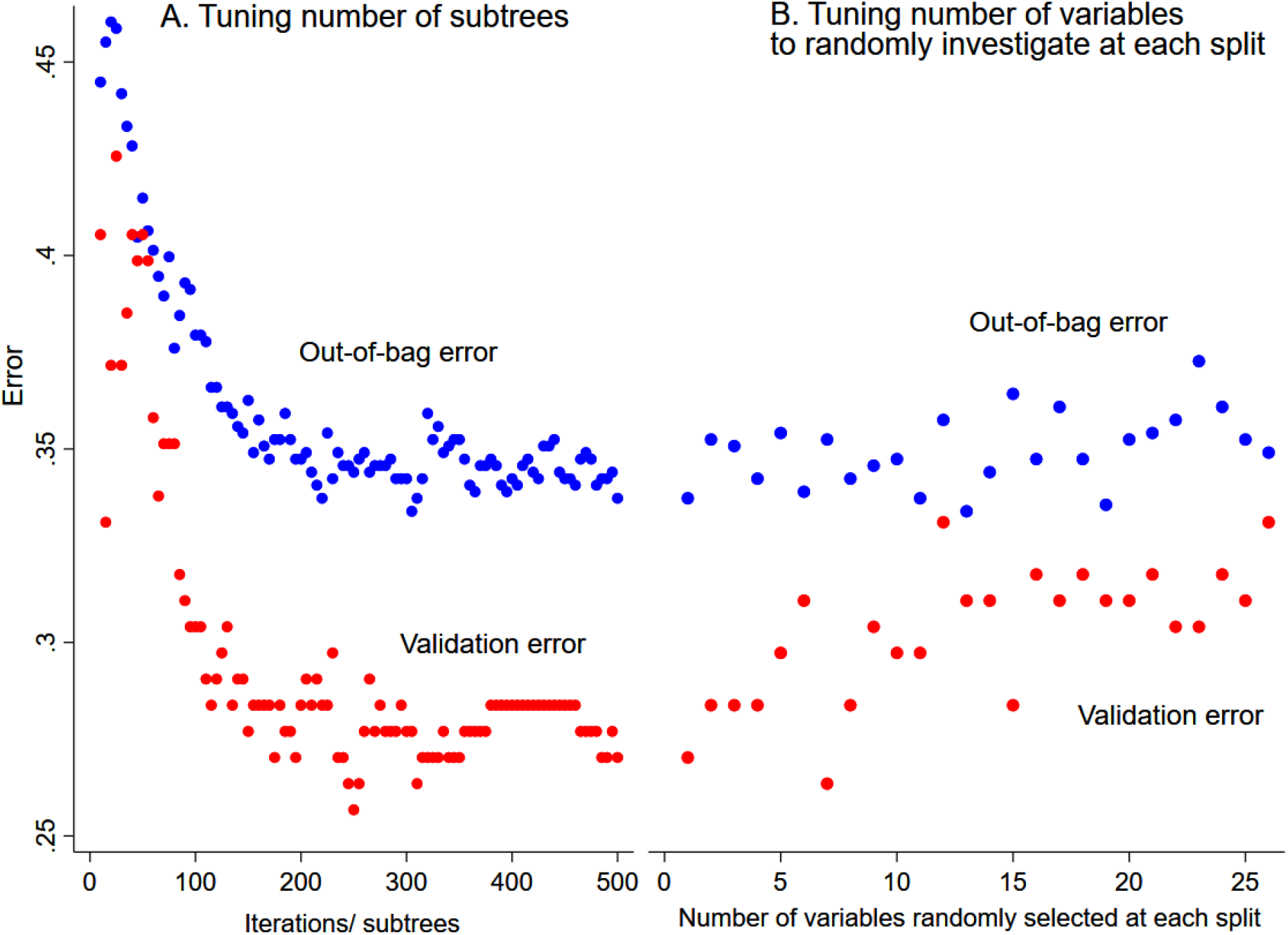
Out-of-bag error and validation error plotted versus (A) number of iterations or subtrees, and (B) number of variables randomly investigated at each split in a random forests model.

**Supplemental Figure 2:**
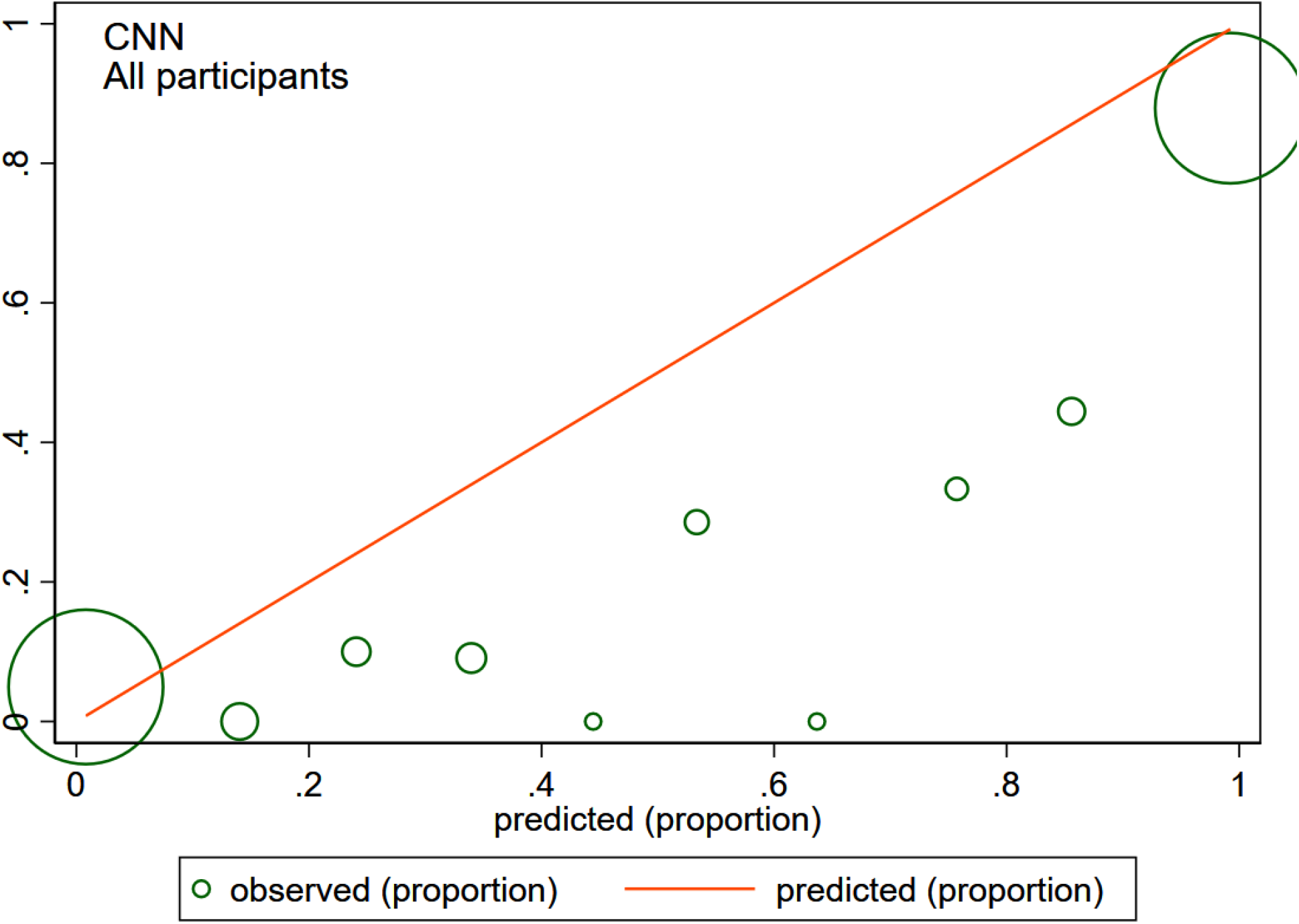
The calibration plot shows the observed and predicted CRT response proportions in convolutional neural network model for all participants. The size of the circles is proportional to the amount of data.

## References

1. Tracy CM, Epstein AE, Darbar D, et al. 2012 ACCF/AHA/HRS Focused Update of the 2008 Guidelines for Device-Based Therapy of Cardiac Rhythm Abnormalities: a report of the American College of Cardiology Foundation/American Heart Association Task Force on Practice Guidelines. Heart Rhythm 2012;9:1737–1753.

2. Chatterjee NA, Singh JP. Cardiac resynchronization therapy: past, present, and future. Heart Fail Clin 2015;11:287–303.

3. Beela AS, Duchenne J, Petrescu A, et al. Sex-specific difference in outcome after cardiac resynchronization therapy. Eur Heart J Cardiovasc Imaging 2019;20:504–511.

4. Linde C, Abraham WT, Gold MR, St. John Sutton M, Ghio S, Daubert C. Randomized Trial of Cardiac Resynchronization in Mildly Symptomatic Heart Failure Patients and in Asymptomatic Patients With Left Ventricular Dysfunction and Previous Heart Failure Symptoms. Journal of the American College of Cardiology 2008;52:1834–1843.

5. Bristow MR, Saxon LA, Boehmer J, et al. Cardiac-resynchronization therapy with or without an implantable defibrillator in advanced chronic heart failure. NEnglJ Med 2004;350:2140–2150.

6. Singh JP, Fan D, Heist EK, et al. Left ventricular lead electrical delay predicts response to cardiac resynchronization therapy. Heart Rhythm 2006;3:1285–1292.

7. Gold MR, Yu Y, Singh JP, et al. Effect of Interventricular Electrical Delay on Atrioventricular Optimization for Cardiac Resynchronization Therapy. Circ Arrhythm Electrophysiol 2018;11:e006055.

8. Cheng A, Gold MR, Waggoner AD, et al. Potential mechanisms underlying the effect of gender on response to cardiac resynchronization therapy: insights from the SMART-AV multicenter trial. Heart Rhythm 2012;9:736–741.

9. Field ME, Yu N, Wold N, Gold MR. Comparison of measures of ventricular delay on cardiac resynchronization therapy response. Heart Rhythm 2020;17:615–620.

10. Deo RC. Machine Learning in Medicine. Circulation 2015;132:1920–1930.

11. Kalscheur MM, Kipp RT, Tattersall MC, et al. Machine Learning Algorithm Predicts Cardiac Resynchronization Therapy Outcomes: Lessons From the COMPANION Trial. Circ Arrhythm Electrophysiol 2018;11:e005499.

12. Tokodi M, Schwertner WR, Kovács A, et al. Machine learning-based mortality prediction of patients undergoing cardiac resynchronization therapy: the SEMMELWEIS-CRT score. European Heart Journal 2020;41:1747–1756.

13. Feeny AK, Rickard J, Patel D, et al. Machine Learning Prediction of Response to Cardiac Resynchronization Therapy: Improvement Versus Current Guidelines. Circ Arrhythm Electrophysiol 2019;12:e007316.

14. Cikes M, Sanchez-Martinez S, Claggett B, et al. Machine learning-based phenogrouping in heart failure to identify responders to cardiac resynchronization therapy. European journal of heart failure 2019;21:74–85.

15. Stein KM, Ellenbogen KA, Gold MR, et al. SmartDelay determined AV optimization: a comparison of AV delay methods used in cardiac resynchronization therapy (SMART-AV): rationale and design. Pacing Clin Electrophysiol 2010;33:54–63.

16. Ellenbogen KA, Gold MR, Meyer TE, et al. Primary results from the SmartDelay determined AV optimization: a comparison to other AV delay methods used in cardiac resynchronization therapy (SMART-AV) trial: a randomized trial comparing empirical, echocardiography-guided, and algorithmic atrioventricular delay programming in cardiac resynchronization therapy. Circulation 2010;122:2660–2668.

17. Matsushita K, Selvin E, Bash LD, Astor BC, Coresh J. Risk implications of the new CKD Epidemiology Collaboration (CKD-EPI) equation compared with the MDRD Study equation for estimated GFR: the Atherosclerosis Risk in Communities (ARIC) Study. Am J Kidney Dis 2010;55:648–659.

18. Tereshchenko LG, Cheng A, Park J, et al. Novel measure of electrical dyssynchrony predicts response in cardiac resynchronization therapy: Results from the SMART-AV Trial. Heart Rhythm 2015;12:2402–2410.

19. Hainmueller J. Entropy Balancing for Causal Effects: A Multivariate Reweighting Method to Produce Balanced Samples in Observational Studies. Political Analysis 2012;20:25–46.

20. Schonlau M, Zou RY. The random forest algorithm for statistical learning. The Stata Journal 2020;20:3–29.

21. BRAIN: Stata module to provide neural network [computer program]. Version 1. Boston: Boston College Department of Economics; 2018.

22. Belloni A, Chen D, Chernozhukov V, Hansen C. Sparse Models and Methods for Optimal Instruments With an Application to Eminent Domain. Econometrica 2012;80:2369–2429.

23. Zou H, Hastie T. Regularization and variable selection via the elastic net. Journal of the Royal Statistical Society: Series B (Statistical Methodology) 2005;67:301–320.

24. Lemeshow S, Hosmer DW, Jr. A review of goodness of fit statistics for use in the development of logistic regression models. Am J Epidemiol 1982;115:92–106.

25. Nattino G, Lemeshow S, Phillips G, Finazzi S, Bertolini G. Assessing the calibration of dichotomous outcome models with the calibration belt. Stata Journal 2017;17:1003–1014.

26. Yancy CW, Jessup M, Bozkurt B, et al. 2013 ACCF/AHA guideline for the management of heart failure: executive summary: a report of the American College of Cardiology Foundation/American Heart Association Task Force on practice guidelines. Circulation 2013;128:1810–1852.

27. Arshad A, Moss AJ, Foster E, et al. Cardiac resynchronization therapy is more effective in women than in men: the MADIT-CRT (Multicenter Automatic Defibrillator Implantation Trial with Cardiac Resynchronization Therapy) trial. J Am CollCardiol 2011;57:813–820.

28. Jamerson D, McNitt S, Polonsky S, Zareba W, Moss A, Tompkins C. Early Procedure-Related Adverse Events by Gender in MADIT-CRT. J Cardiovasc Electrophysiol 2014.

29. Haq K, Cao J, Tereshchenko LG. Characteristics of Cardiac Memory in Patients with Implanted Cardioverter Defibrillator: the CAMI study. medRxiv 2019; https://doi.org/10.1101/19005181.

30. Waks JW, Perez-Alday EA, Tereshchenko LG. Understanding Mechanisms of Cardiac Resynchronization Therapy Response to Improve Patient Selection and Outcomes. Circ Arrhythm Electrophysiol 2018;11:e006290.

31. Rahman QA, Tereshchenko LG, Kongkatong M, Abraham T, Abraham MR, Shatkay H. Utilizing ECG-Based Heartbeat Classification for Hypertrophic Cardiomyopathy Identification. IEEE Trans Nanobioscience 2015;14:505–512.

32. Hu SY, Santus E, Forsyth AW, et al. Can machine learning improve patient selection for cardiac resynchronization therapy? PloS one 2019;14:e0222397.

33. Tereshchenko LG, Henrikson CA, Stempniewicz P, Han L, Berger RD. Antiarrhythmic Effect of Reverse Electrical Remodeling Associated with Cardiac Resynchronization Therapy. PACE-Pacing Clin Electrophysiol 2011;34:357–364.

34. Tereshchenko LG, Henrikson CA, Berger RD. Strong coherence between heart rate variability and intracardiac repolarization lability during biventricular pacing is associated with reverse electrical remodeling of the native conduction and improved outcome. Journal of Electrocardiology 2011;44:713–717.

35. Jacobsson J, Borgquist R, Reitan C, et al. Usefulness of the Sum Absolute QRST Integral to Predict Outcomes in Patients Receiving Cardiac Resynchronization Therapy. Am J Cardiol 2016;118:389–395.

